# The TuberXpert project protocol: Towards a Clinical Decision Support System for therapeutic anti-tuberculosis medical drugs monitoring in Tanzania

**DOI:** 10.1101/2023.12.22.23300427

**Authors:** Yann Thoma, Annie E. Cathignol, Yuan J. Pétermann, Margaretha L. Sariko, Bibie Said, Chantal Csajka, Monia Guidi, Stellah G. Mpagama

**Author notes:** Co-senior authors (Monia Guidi, Stellah G. Mpagama).

## Abstract

**Introduction:** The End Tuberculosis (TB) Strategy requires a novel patient treatment approach contrary to the “one-size fits all” model. It is well known that each patient’s physiology is different and leads to various rates of drug elimination. Therapeutic Drug Monitoring (TDM) offers a way to manage drug dosage adaptation but requires trained pharmacologists, which is scarce in resource-limited settings. We, therefore, aim to create an unprecedented Clinical Decision Support System (CDSS) that will offer a printable report containing advice for the field clinicians to guide the adaptation of TB treatment depending on the patient.

**Methods and analysis:** A population pharmacokinetic model for rifampicin will be developed and thoroughly validated, before being implemented into Tucuxi, an existing Model Informed Precision Dosing software. A cross-sectional study will be conducted to define the best way to display information to clinicians. In addition, a pragmatic prospective study will focus on a decision tree that will be implemented as a CDSS. Expert pharmacologists will validate the CDSS, and, finally, field implementation in Tanzania will occur coupled with a prospective study to assess clinicians’ adherence to the CDSS recommendations.

**Ethics and dissemination:** This is a game-changing transdisciplinary project combining technology and pharmacometrics to enable appropriate dosages of anti-TB drugs in TB patients at various levels of the healthcare delivery system in TB-endemic settings. The project is part of the Adaptive Diseases control Expert Programme in Tanzania, which has been approved at the local health research committee serving Kibong’oto Infectious Diseases Hospital (KIDH) and National Health Research Committee with reference numbers KNCHREC003 and NIMR/HQ/R.8a/Vol.IX/2988, respectively. Furthermore, the Ministries of Health and Regional Administrative & Local Government Authority have endorsed the implementation of this protocol. Dissemination will be done through scientific publications, conferences, and local press in Tanzania. Social media will also be used to gain more visibility.

**Strengths and limitations of this study:**

- To our knowledge, this is the first study to investigate the application of CDSS technology at varying healthcare delivery systems levels to guide TDM in TB patients in TB-endemic settings.
- Routine implementation of TDM-CDSS, particularly for rifampicin, a backbone for TB treatment, is expected to transform TB’s clinical management in resource-limited settings.
- Anti-TB dosage optimization will improve treatment outcomes of patients who would otherwise succumb or develop drug-resistant TB because of sub-optimal drug exposure. This will considerably contribute to the End TB strategy, particularly with arduous forms of TB with either Human Immunodeficiency Virus (HIV) co-infection or coexistent Diabetes Mellitus (DM) or malnutrition.
- Clinicians could also use the CDSS decision tree without access to IT infrastructure.
- A lack of computer infrastructure in health facilities may prevent implementing a centralized system in resource-limited countries.

## 1 Introduction

Even though tuberculosis (TB) is curable, it remains one of the leading causes of death from a single bacterial infection worldwide. The World Health Assembly declared the end of TB with a drastic reduction in death and incidence; however, the decrease is at a sub-optimal pace, only 11% and 9.2%, contrary to at least 20% and 35% by 2020, respectively [1] [2]. Sub-Saharan Africa contributes around 25% of the global TB burden, with a higher death rate than other World Health Organisation (WHO) regions at 22% [3]. Significant and commonly co-existing comorbidities in TB patients, which include Human Immunodeficiency Virus (HIV) coinfection [4], enteric pathogens [5], malnutrition [6], and diabetes mellitus (DB) [7], increase the risk of unfavorable treatment outcomes. This is primarily driven by incomplete adherence and pharmacokinetic variability of first-line antitubercular (anti-TB) drugs, leading to insufficient circulating drug exposure and development of drug-resistant TB or excessive exposure and toxicity, resulting in treatment interruption.

Precision dosage of medicines based on drug concentration monitoring is an essential patient-centered approach for optimizing adherence and efficacy and preventing adverse effects. There are available tools for helping pharmacologists with the dosage adaptation process. However, they are difficult to use for non-specialists and would benefit from automation. Real Clinical Decision Support Systems (CDSS), which have the potential to assist practitioners, not necessarily pharmacologists, in the dosage adaptation of anti-TB drugs in TB patients, are yet to be available in TB endemic settings. Deployment of such an advanced system will encourage the spread of a dosage-individualization culture among practitioners not specialized in pharmacology while bridging the gap by adapting those novel technologies for optimizing TB care [8].

In that context, as part of the Adaptive Diseases control Expert Programme in Tanzania, the TuberXpert project aims to develop an automated CDSS to help practitioners with the dosage adaptation of rifampicin. Being one of the essential medical drugs targeting TB, rifampicin is known for its significant pharmacokinetic variability and frequent suboptimal blood exposure [9].

This protocol shows the field application of such an automated CDSS for the dosage adaptation of rifampicin in Tanzania with underpinned applied research questions to address scientific implementation for sustainability.

## 2 Methods and analysis

An automated CDSS software requires an embedded population pharmacokinetic (popPK) model for rifampicin to predict drug concentrations and propose meaningful adjustments correctly. The software will be based on the core computing engine of Tucuxi [10, 11], a model-informed precision dosing software for Bayesian forecasting already developed that helps clinicians adapt medical drug dosages. This software computations are based on the popPK models implemented within the software and the model-identified influential patient covariates, potential drug concentration measurements, and specific drug PK targets validated by TDM experts for dose optimization. The CDSS will be implemented in five phases, as shown in Figure 1. In brief, the first phase includes developing an appropriate popPK model for rifampicin for Tanzanian patients and implementing it within Tucuxi. The second phase will optimize reporting relevant information to practitioners for drug dosage adjustment. The third phase will automate the delivery of the report in line with the measurement of the drug concentration [12]. The fourth phase will validate the system, and the final step will implement the system in the field. Details of each stage and associated research questions are summarized from 2.1 through 2.5.

**Figure 1.**
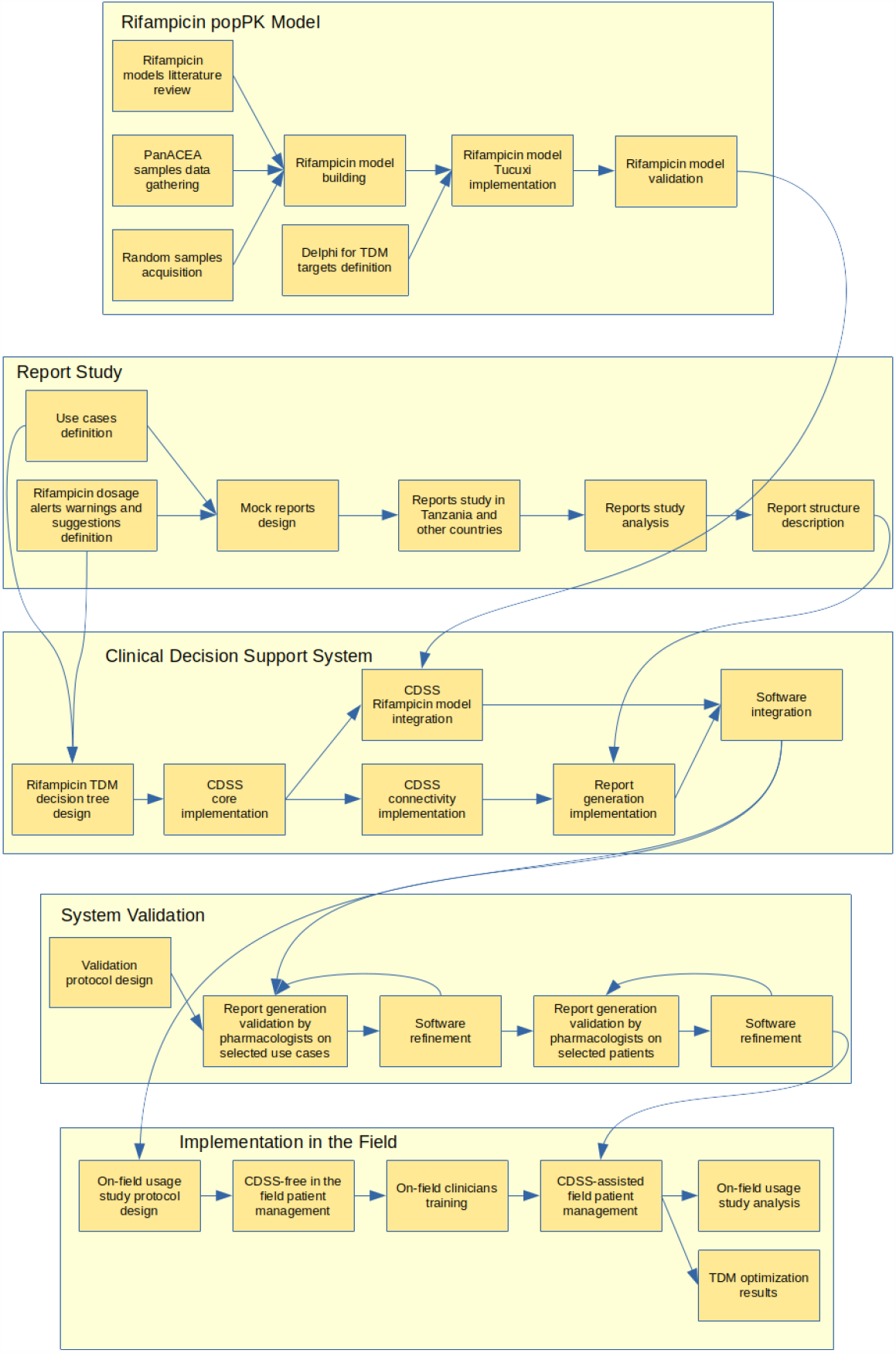
TuberXpert protocol PERT diagram, decomposed into five phases.

### 2.1 Model selection, development, and calibration

Predicting a concentration of a medical drug in blood for dosage adjustment using Tucuxi crucially requires the selection of a popPK model, as the latter determines the ability of the software to propose the correct dosage regimen for each patient. An extensive literature search will be conducted to identify the published popPK models of rifampicin in TB patients [13, 14, 15], developed using NONMEM [16] or Monolix [17], both widely used computer programs for parametric popPK analyses. Models describing rifampicin PK in TB patients with the previously mentioned comorbidities, i.e., DM, malnutrition and HIV, will be included to investigate their impact on the anti-TB drug PK. The number of participants in the studies, as well as that of available observations distributed over time, will be considered. The description of the model development steps (e.g., structural and statistical considerations, inclusion of covariates) will be carefully reviewed, and the study results will be judged using standard procedures in population analysis (e.g., diagnostic goodness-of-fit plots, type of validation) to ensure the quality of the model and the completeness of the information provided. The available models will be ranked according to a cumulative score based on the previous elements, and the best one will be selected for implementation. Because of the autoinduction and the non-linear processes underlying rifampicin absorption and elimination, we expect a complex non-linear popPK model to characterize its PK best (which will require software updates to encompass specific kinetics). If the best-identified model fails in capturing essential PK characteristics of rifampicin, the opportunity to use a model-based meta-analysis (MBMA) approach will be investigated to build a comprehensive popPK model using all available pertinent literature. Model refinement will be attempted if the selected model proves to be computationally prohibitive for its implementation in Tucuxi, verifying that the simplified model’s prediction for the targeted exposures is comparable to the original ones. Whatever the choice made for this step, the model will undergo formal validation and adjustment to Tanzanian patients using rifampicin concentrations available through the panACEA network and those collected in Tanzania within this project. To achieve this aim, a prospective study will be conducted on 50 TB patients (including those with HIV co-infection, coexistent DM, or malnutrition) followed at the Kibong’oto Infectious Diseases Hospital (KIDH) or in neighborhood clinics during the early stage of the TuberXpert project. These patients will provide three samples randomly on two separate occasions, i.e., at the treatment start and approximately after one week of treatment. We will elaborate and then strictly follow a study protocol. We will also conduct a study comparing Dried Blood Spot (DBS) analysis [12] to HPLC-UV lab analysis [18] on these samples. Finally, we will conduct a Delphi study, as suggested in [19], to define the optimal therapeutic target, as this is critical for therapy individualization and there is currently no definitive consensus for rifampicin (various propositions in [13, 20, 21, 22]).

A further essential preliminary validation step for the successful use of Tucuxi in therapy individualization is to ensure that the dosage regimen propositions made by the software agree with those of experienced pharmacologists used to perform Therapeutic Drug Monitoring (TDM) in their clinical practice. Pharmacologists of the service of clinical pharmacology of the University Hospital Center of Vaud at Lausanne (CHUV) and clinicians at KIDH will be in charge of such clinical validation of the software using selected patients of the current study. Data on rifampicin dosage history and concentrations, administration and sampling times, and all necessary clinical information on the selected patients will be collected. An experienced pharmacologist will choose the most appropriate dosage regimen to achieve the target according to the patient clinical status among those proposed by the software using the Graphical User Interface of Tucuxi. The correctness of the forecasted dosing schedule will be validated against the dosage recommendations made by the TDM experts at CHUV and clinicians at KIDH. After this vital validation step, the model will be made available within the standalone Tucuxi software, allowing users to exploit this new model.

The research question components are:

◼ What is the appropriate popPK model that includes the structural and statistical considerations, as well as the influential individual factors, to be incorporated in Tucuxi for optimizing rifampicin dosage in TB patients?
◼ What is the Tucuxi-guided rifampicin dosage adjustment accuracy for anti-TB therapy individualization against experienced pharmacologists in the TDM clinical practice?

### 2.2 Report design specifications

Report generation, specifically the report structure and visual content, is essential to help clinicians with decision-making. It shall be easily understandable by non experienced TDM pharmacologists. The displayed information must be carefully selected and structured to not lose the prescriber with a too complex presentation and terminology [23]. It should contain the appropriate information presented visually. In addition, alerts shall be issued, if necessary, in a form following the I-MeDeSa principles [24].

The definition of the report layout will be elaborated through the following process, depicted in Figure 2. We will prepare mock reports showing various layouts based on use cases. The designs will be done after reviewing the literature to identify good practices. We will then, in Tanzania, expose a cohort of clinicians to these reports and collect their input through semi-structured interviews. We expect to reach ten clinicians from different facilities, in Tanzania. We also aim to ask ten non-Tanzanian clinicians to get helpful information about varying preferences regarding information representation depending on culture or background. Thus, pharmacologists with a strong expertise in dosage adjustment will be included to get their specific needs compared to general medical doctors. We will improve the report’s rendering over two cycles of interviews, with thorough attention to the clinicians’ feedback. The output of this study will allow for the design of the final report. This study will also illustrate the differences between TDM expert pharmacologists and general medical doctors and, hopefully, between other countries and Tanzania for report design acceptance.

**Figure 2.**
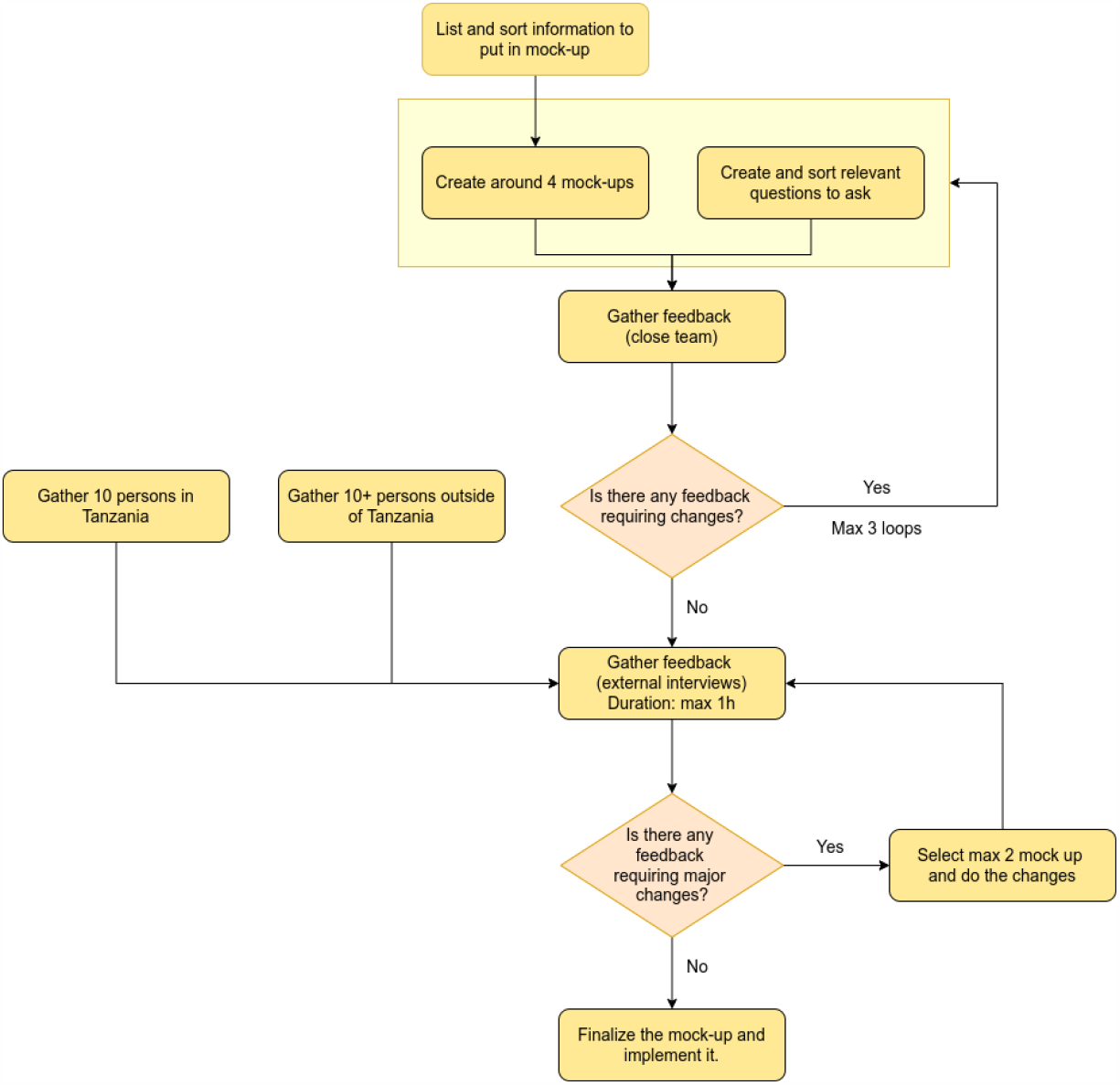
Report design study protocol.

The research question components are:

◼ What information and how this information shall be displayed to clinicians for an efficient decision-making process concerning dosing regimen?
◼ What are the differences in terms of expectations about reports data and design among expert pharmacologists and medical doctors, both in Tanzania and Western world?

### 2.3 Clinical Decision Support System

Tucuxi offers a graphical user interface, graphs with drug concentration predictions surrounded by percentiles, and dosage adaptation propositions. The software uses Bayesian inference to find the most likely individual parameters integrating information from the underlying popPK model and the observations obtained on the patient. Currently, the user has to decide by himself/herself on the dosage adjustment among those proposed by the software and write some sentences to fill up a report. Experienced pharmacologists do this manually, but this practice requires particular skills related to this specialized medical discipline. Clinicians who make final decisions about the drug dosage rely on monographs, experience, or a consultation with a pharmacologist for dosage adaptation. In that context, a CDSS can bring much needed information to decision-makers [25] in locations where expert pharmacologists are not readily available.

The CDSS that will be developed in TuberXpert will offer various features: (1) assess the expectedness (likelihood) of a drug concentration result, taking into account the patient’s characteristics; (2) assess the adequateness (target attainment) of the current dosage; (3) propose a dosage adjustment if required; (4) present clear and meaningful messages within the report to help the clinician with the decision-making process; and (5) generate alerts when data seems suspicious or erroneous.

While the current Tucuxi software allows the necessary computations, this project will add a new level to the computing engine. Research is needed to ensure the system is extensible enough to address other medical drugs without colossal refactoring. Therefore, a perfect balance between genericity and TB/rifampicin-specific features has to be found. The system must also be easily integrated into existing IT systems thanks to a generic interface. It will also need to be highly reliable and deployable in the field.

Common and uncommon use cases (e.g., type of patient, under/over drug exposure, standard/strange observed drug level) will be identified in both Switzerland and Tanzania, to increase case heterogeneity. Indeed, the TB patients treated in Switzerland are usually exceptional cases compared to the daily routine TB patients in Tanzania. Artificial use cases will complete this panel to cover as many borderline cases as possible. We will carefully analyze every patient and use case scenario to design a formal decision tree [26] that will lead to software development. This decision tree will be required to create the software and will be published as a guide for clinicians doing the job by hand. Special attention will be given to appropriate corrective actions [27]. For instance, if a measured drug level appears out of range, an alert shall identify either a potential error in the data or a lack of adherence whenever such sources of bias can be identified with sufficient certainty. In less clear situations, an estimation of the likelihood of common artifacts vs a definite individual PK alteration in the patient should be issued.

The report to be generated shall display proper values and graphs and offer readable sentences. In close relation with the decision tree, a set of standard customizable sentences will have to be defined. They will then be integrated to offer medically meaningful reports as if a human professional had written them. These sentences will be related to TB and rifampicin. Still, the software will be developed keeping in mind that it should be straightforward to define new sentences for other diseases and medicinal drugs. Thanks to established configurations, the core components will be generic, and a parametrization will enable repurposing as efficiently as possible.

We will conduct a short survey in Tanzania to address the availability of computer infrastructures, printers, and connectivity in clinics and at the doctor’s desk. We also will ensure that the system can be connected to the laboratory information system that manages the transmission of drug concentration measurement results. In addition, having a version of the CDSS where computations are running remotely could be an option if the clinicians in the field do not own computers with enough computing power. If this approach is required, the patient’s data could stay on the clinician’s computer to ensure data privacy.

The research question components are:

▪ What data processing leads to the most accurate suggestions based on the available data of a specific patient?
▪ What setup is the most appropriate for rural clinics where access to computer infrastructures is not ideal?

### 2.4 System validation

TuberXpert will then be validated after the CDSS and report generation are completed, considering the rifampicin popPK model has already been validated and calibrated to the target population. We will assess the correctness of the generated reports based on use cases designed within the project among patients on rifampicin followed at CHUV and on the data gathered in Tanzania (retrospective data of patients from CHUV and 25 from Tanzania, i.e., randomly selected 50% among hospital/study patients). CHUV patients necessitating a TDM intervention often present an overly complex clinical situation, allowing for report-generation testing in extraordinary cases. Experienced pharmacologists will generate a manual report using the Graphical User Interface of Tucuxi to individualize the therapy of the selected patients. Such a “standard” document will contain all the essential information to justify the chosen dosage adjustment, including the patient clinical situation, data non-reliability, or inconsistencies. It will serve as the gold standard for the TuberXpert report, which will be compared to it to identify any discrepancies. We will verify the correctness of the provided guidance and the presence of all the necessary clinical information, justifications, and warnings. This step will also allow for the refinement of the report to guarantee the reliability of the information for the final user, i.e., a clinician without strong TDM expertise. Before conducting this project phase, we will elaborate a detailed protocol to ensure the most reliable validation.

After this first validation step, we will update the CDSS and the report generation based on the study results and specifically on the feedback of the pharmacologists. Indeed, the CDSS development will be driven by the use cases defined in the project beginning, while the validation will be conducted on another set of retrospective data. As such, some adjustments will likely be required.

After the software update, we will conduct a second validation, following the same protocol but on another set of patients (25 from Tanzania and 15 from CHUV). This second step should end with very few differences between the pharmacologists’ outputs and the generated reports, and we only expect minor software modifications to be necessary after that. Finally, we will again run TuberXpert against all the use cases to validate it against the pharmacologists’ choices made during the two validation steps.

The research question component is:

▪ Is the CDSS developed reliable enough in comparison with a human pharmacologist?

### 2.5 Implementation in the field

The last project phase consists of the CDSS implementation in the field and a prospective study following a strict protocol that will be elaborated according to Tanzania laws on human research. The goal will be to observe the relevance of the generated reports for the clinicians in Tanzania. KIDH will select a cohort of clinicians actively involved in TB treatment with rifampicin that will identify patients to enroll in the study. This step will assess the benefit of the CDSS use versus standard practice in Tanzania. Thanks to the existing network of KIDH, we expect to have at least 30 patients in the cohort treated in 10 facilities in the field.

Before introducing TuberXpert to the clinicians, the selected panel will follow their patients according to standard clinical management while collecting all the needed information for a TDM procedure using a dedicated form that will be elaborated according to CDSS specificities. In addition, the possible dosage adjustments, the patient’s health status evolution (i.e., clinical features and pragmatic microbiology parameters), and all other relevant data about the patients will be collected. Blood analysis will be implemented and centralized at Kilimanjaro Clinical Research Institute (KCRI) or at KIDH. The KIDH will use this information to feed the CDSS to determine the correspondence between the CDSS output and the clinicians’ decisions. The CDSS will be used in this phase without interfering with the patient’s treatment. Then, the consortium will train the clinicians in the field about CDSS usage and report interpretation. This training will not only be beneficial for the current project but also for spreading the TDM culture throughout Tanzania.

After the training session, the CDSS will be available at the clinics participating in the study. We will deploy the software in facilities where such deployment is feasible, in a setup that still has to be defined, also setting up a direct communication channel with the Swiss team to help the clinicians with any questions regarding software usage or TDM in general. More clinicians will be involved in this second phase, including but not limited to those who participated in the previous non-CDSS phase, as participation in the first phase is not mandatory to be part of the second one. The study protocol will be written during the project and will strongly focus on the clinical relevance of the CDSS and the acceptance of the software tool [28]. The clinicians will be asked to collect the same information as in the first implementation phase, including clinical features during the patients’ follow-up and pragmatic microbiology parameters. Likewise, we will establish the baseline information of the safety of TDM by observing and assessing patients’ clinical evolution between the two phases, which will subsequently offer the opportunity to determine the overall benefit of CDSS-assisted *vs* standard patient management in Tanzania. This second part will also help identify the cases where TuberXpert has influenced the clinicians, why they would not be keen to follow the suggested regimens, and if the outcome of the dosage adjustment has been positive for the patient. If clinicians scarcely follow the CDSS suggestions, we will investigate the barriers and bottlenecks that hinder its implementation to increase the accessibility of benefits of the TuberXpert technology. The analysis will then be performed by comparing the subgroup of clinicians that followed the CDSS recommendations vs those who were reluctant and included in the first phase to assess the CDSS benefits.

The research question component is:

▪ Are clinicians keen to adhere to the suggestions of an automated CDSS?

## 3 Conclusion

This game-changing transdisciplinary project combines technology and pharmacometrics to enable appropriate dosages of anti-TB drugs in TB patients at various levels of the healthcare system in TB-endemic settings. TuberXpert thus provides an unprecedent opportunity to optimize and individualize the clinical management of TB patients in resource-limited settings. Moreover, TDM tends to be increasingly recommended, particularly for treating multidrug-resistant TB with second-line chemotherapeutic agents, when achieving effective and safe concentration exposure is of vital importance [29]. TuberXpert could be adapted beyond rifampicin and holds promise far beyond tuberculosis alone, with indirect benefits in other therapeutic areas such as HIV, cancer, etc.

## Data Availability

This paper presents a project at its beginning, so no data is yet available.

## 4 Patients and Public Involvement

The development of this protocol aligns with the patient-centered care approach [30]. This project was designed based on the results of a series of TB research studies in Tanzania that showed individual TB patients differ and may need different treatment strategies and duration due to variations of disease severity and comorbidities [31, 32, 33, 34, 35]. Findings from the described project will be shared with TB patients’ organizations for further refinement before subsequently contributing to shaping the agenda of effective integration of communicable and non-communicable diseases for policymakers.

## 5 Ethics and dissemination

The patient enrolment has been approved at the local health research committee serving Kibong’oto Infectious Diseases Hospital and National Health Research Committee with reference numbers KNCHREC003 and NIMR/HQ/R.8a/Vol.IX/2988, respectively. Furthermore, the Ministries of Health and Regional Administrative & Local Government Authority have endorsed implementing this protocol. Dissemination will be done mainly through scientific publications for all project parts. Social media will also be used to gain more visibility. Finally, spreading the usage of the system is a goal envisioned at the end of the project.

## Contributors

YT, MG, and SGM conceptualized, designed, and supervised the research proposal. YT wrote the manuscript with input from all the authors. YT is leading the project, specifically the software part of it. MG is leading the popPK rifampicin model development and coordinating the CHUV team. SGM is leading the study in the field. AC is in charge of conducting the report study and the CDSS development. YP is developing the rifampicin popPK model and is working on the TDM implementation. BS is in charge of the studies in the field (enrolment of patients and implementation). MS is responsible for the lab analysis. CC is co-supervising AC and YP PhDs. All authors have approved the final version and agreed to be accountable for all aspects of the work related to accuracy and integrity.

### Funding

This work is funded by the Swiss National Science Foundation, under grant IZSTZ0_208544.

